# Multi-organ impairment in low-risk individuals with long COVID

**DOI:** 10.1101/2020.10.14.20212555

**Authors:** Andrea Dennis, Malgorzata Wamil, Sandeep Kapur, Johann Alberts, Andrew D. Badley, Gustav Anton Decker, Stacey A Rizza, Rajarshi Banerjee, Amitava Banerjee, On behalf of the COVERSCAN study investigators

## Abstract

**Background:** Severe acute respiratory syndrome-coronavirus 2 (SARS-CoV-2) infection has disproportionately affected older individuals and those with underlying medical conditions. Research has focused on short-term outcomes in hospital, and single organ involvement. Consequently, impact of long COVID (persistent symptoms three months post-infection) across multiple organs in low-risk individuals is yet to be assessed.

**Methods:** An ongoing prospective, longitudinal, two-centre, observational study was performed in individuals symptomatic after recovery from acute SARS-CoV-2 infection. Symptoms and organ function (heart, lungs, kidneys, liver, pancreas, spleen) were assessed by standardised questionnaires (EQ-5D-5L, Dyspnoea-12), blood investigations and quantitative magnetic resonance imaging, defining single and multi-organ impairment by consensus definitions.

**Findings:** Between April and September 2020, 201 individuals (mean age 44 (SD 11.0) years, 70% female, 87% white, 31% healthcare workers) completed assessments following SARS-CoV-2 infection (median 140, IQR 105-160 days after initial symptoms). The prevalence of pre-existing conditions (obesity: 20%, hypertension: 6%; diabetes: 2%; heart disease: 4%) was low, and only 18% of individuals had been hospitalised with COVID-19. Fatigue (98%), muscle aches (88%), breathlessness (87%), and headaches (83%) were the most frequently reported symptoms. Ongoing cardiorespiratory (92%) and gastrointestinal (73%) symptoms were common, and 42% of individuals had ten or more symptoms.

There was evidence of mild organ impairment in heart (32%), lungs (33%), kidneys (12%), liver (10%), pancreas (17%), and spleen (6%). Single (66%) and multi-organ (25%) impairment was observed, and was significantly associated with risk of prior COVID-19 hospitalisation (p<0.05).

**Interpretation:** In a young, low-risk population with ongoing symptoms, almost 70% of individuals have impairment in one or more organs four months after initial symptoms of SARS-CoV-2 infection. There are implications not only for burden of long COVID but also public health approaches which have assumed low risk in young people with no comorbidities.

**Funding:** This work was supported by the UK’s National Consortium of Intelligent Medical Imaging through the Industry Strategy Challenge Fund, Innovate UK Grant 104688, and also through the European Union’s Horizon 2020 research and innovation programme under grant agreement No 719445.

## Introduction

Early in the COVID-19 pandemic, research and clinical interest in SARS-CoV-2 (Severe acute respiratory syndrome-coronavirus 2)-induced organ damage was predominantly focused on the respiratory system(1). There have been indirect effects on other organ systems and disease processes, such as cardiovascular diseases and cancers, through changes in health systems or behaviours of patients and health professionals(2-4). In addition, beyond an acute systemic inflammatory response, evidence for direct COVID-19-related effects on multiple organs is accumulating, with potential long-term impacts for individuals as well as health systems(5-8). However, no study to-date has included detailed characterisation of all major organ systems following SARS-CoV-2 infection.

COVID-19 represents a convergence of an infectious disease, under-treated non-communicable diseases and social determinants of health, as a “syndemic”(9). Pre-existing non-communicable diseases and risk factors are important predictors of poor COVID-19 outcomes, whether intensive care admissions or mortality(2). Research has focused on the acute phase of SARS-CoV-2 infection, in hospitalised patients, and on individuals that have died from COVID-19(10-12). It is clear that COVID-19 can have longer multiple symptoms and long-term effects(13), but “long-COVID” is yet to be fully defined(14-15), partly due to lack of understanding of medium- and long-term pathophysiology across organ systems.

Long COVID in low-risk individuals, who represent up to 80% of the population(2), has public health importance in terms of burden of disease and healthcare utilisation, and therefore has urgent policy relevance across countries. However, in the UK, government policies have emphasised excess risk of mortality in moderate- and high-risk conditions, including “shielding”(2) and commissioning of a risk calculator to identify those at highest risk of COVID-19 severity and mortality(16). As the pandemic progresses, there is growing concern regarding prolonged isolation strategies for people with vulnerable conditions and at highest risk of severe COVID-19 outcomes(17). These approaches have assumed low risk of SARS-CoV-2 infection in younger individuals without underlying conditions, based on their low excess mortality, but without knowledge of the chronic pulmonary and extrapulmonary effects of COVID-19.

In order to better understand the long-term impact of COVID-19 and ultimately inform preventive measures at health system level, we performed a pragmatic, prospective study in low-risk individuals with symptom assessment, multi-organ magnetic resonance imaging (MRI) and blood investigations for inflammatory markers at three months post-COVID-19 diagnosis.

## Methods

### Patient population and study design

In an ongoing, prospective study, 201 participants were enrolled at two UK sites (Perspectum, Oxford and Mayo Clinic Healthcare, London) between April 2020 and August 2020 and completed baseline assessment by 14 September 2020 (**Figure 1**). Participants were eligible for enrolment if they tested positive by the oro/nasopharyngeal throat swab for SARS-CoV-2 by reverse-transcriptase-polymerase-chain reaction (n=62), a positive antibody test (n=63), or had typical symptoms and were determined to have COVID-19 by two independent clinicians (n=73). Exclusion criteria were symptoms of active respiratory viral infection (temperature >37.8°C or three or more episodes of coughing in 24 hours); discharged from hospital in the last 7 days; and contraindications to MRI, including implanted pacemakers, defibrillators, other metallic implanted devices; claustrophobia. The study protocol was approved by a UK ethics committee (20/SC/0185), registered (https://clinicaltrials.gov/ct2/show/NCT04369807) and all patients gave written informed consent.

**Figure 1:**
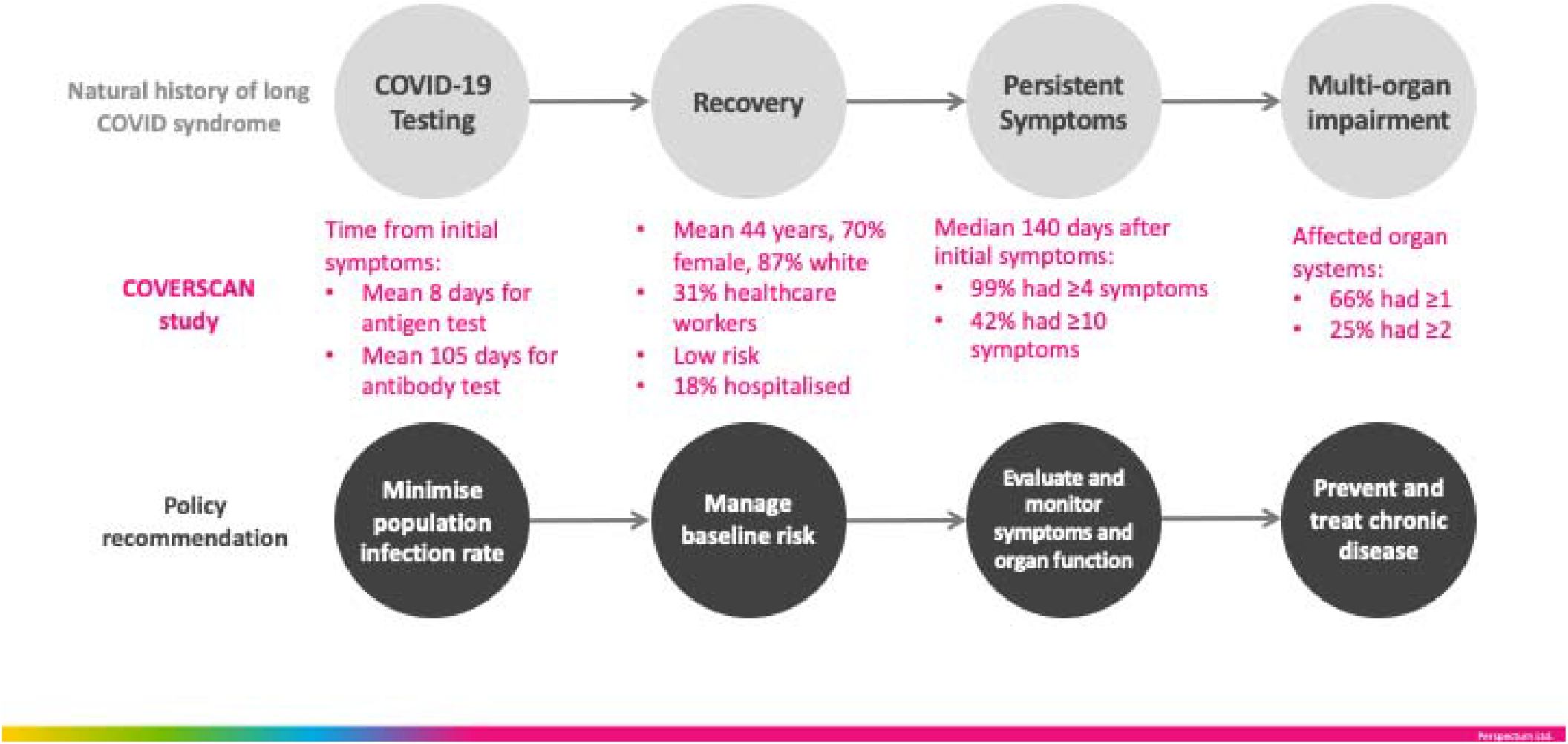
Natural history of long COVID, the COVERSCAN study in low-risk individuals (n=201) and policy recommendations.

### To assess the burden of multi-organ involvement after SARS-CoV2 infection

Organ function was assessed by patient-reported validated questionnaires, fasting blood investigations (as listed below) and multi-organ MRI. MRI was the chosen imaging modality (as in UK Biobank) because it is: (1) safe, with no radiation exposure, no need for intravenous contrast, minimal contact with the radiographer; (2) quantitative, repeatable and robust, with >95% acquisition and image processing success rate; (3) informative through a repository of digital data which can be shared in the research community for independent analysis and research; (4) rapid and scalable, i.e. a 35-minute scan can phenotype the lung, heart, kidney, liver, pancreas and spleen. At time of MRI, we completed (i) questionnaires for quality of life(EQ-5D-5L(18)), addressing mobility, self-care, usual activity, pain and anxiety, and breathlessness (Dyspnoea-12(19)) and (ii) full blood count, serum biochemistry (sodium, chloride, bicarbonate, urea, creatinine, bilirubin, alkaline phosphatase, aspartate transferase, alanine transferase, lactate dehydrogenase, creatinine kinase, gamma-glutamyl transpeptidase, total protein, albumin, globulin, calcium, magnesium, phosphate, uric acid, fasting triglycerides, cholesterol (total, HDL, LDL), iron, iron-binding capacity (unsaturated and total) and inflammatory markers (erythrocyte sedimentation rate, ESR; high sensitivity-C-Reactive Protein, CRP) (TDL laboratories, London).

### Magnetic Resonance Image Analysis

Multi-organ MRI data were collected at both□study sites (Oxford:□MAGNETOM Aera 1.5T, Mayo Healthcare London:□MAGNETOM Vida 3T; both from□Siemens Healthcare Erlangen, Germany). The COVERSCAN Multiparametic MRI assessment typically required 35mins per patient, including lungs, heart, liver, pancreas, kidneys and spleen by standardised methodology (**Supplementary methods**).

### Definition of organ impairment

MRI-derived measurements from the heart, lungs, kidney, liver, pancreas and spleen were compared with established reference ranges (**Table S1**) to determine impairment for each organ. An individual organ was classified as impaired if at least one of the metrics calculated for that organ was outside the reference range. Excessive organ fat was not considered as an indicator of impairment on the assumption that this was likely pre-existing and thus treated separately. Organ impairment was defined for each metric according to established cut-offs (Table S1) and was grouped by evidence of: borderline or low ejection fraction and evidence of myocarditis in the heart; reduced pulmonary dynamic measurements in the lungs; elevated cortical T1 in the kidneys; borderline or definite inflammation in the liver and pancreas; and splenomegaly from spleen length.

### Statistical analysis

All statistical analyses were performed using R software (version 3.6.1) with a p-value less than 0.05 considered statistically significant. Descriptive statistics were used to summarise baseline participant characteristics. Mean and standard deviation (SD) were used to describe normally distributed-continuous variables, median with interquartile range (IQR) for non-normally distributed, and frequency and percentage for categorical variables.

Mean difference in quantitative organ metrics between hospitalised versus not hospitalised were compared using the Wilcoxon test, and difference in the counts of the binary outcomes of those with evidence of organ abnormalities compared using Fisher’s test. Multi-organ impairment was defined as impairment to ≥2 organs. Associations between multi-organ impairment and symptoms, comorbidities and pre-existing risk factors were assessed using Spearman’s correlation. Based on the observed differences between hospitalised and non-hospitalised groups, multivariate logistic regression models were used to assess risk factors for COVID-19 hospitalisation.

## Results

The mean age was 44.0 (SD: 11.0) years. 70% of individuals were female, 87% were white, 31% were healthcare workers, 18% had been hospitalised with COVID-19. Assessment (symptoms, blood and MRI) was a median 140 (IQR 105-160) days after initial symptoms. Relevant past medical history included smoking (3%), asthma (18%), obesity (20%), hypertension (6%), diabetes (2%) and heart disease (4%). The hospitalised group were older (p=0.001), had a higher proportion of non-white participants (p=0.038), and were more likely to report ‘inability to walk’ (p=0.01) than non-hospitalised individuals. There were no other significant differences between risk factors or symptoms reported between the groups. The most commonly reported on-going symptoms (regardless of hospitalisation status) were fatigue (98%), muscle ache (88%), shortness of breath (87%) and headache (83%) (**Table 1, Figure 2(a)**). Ongoing cardiorespiratory (92%) and gastrointestinal (73%) symptoms were common. 99% of individuals had four or more and 42% had ten or more symptoms. 52% of patients reported persistent moderate problems undertaking usual activities (level 3 or greater in the relevant EQ-5D-5L question). 20% reported Dyspnoea-12 ≥15 (equivalent to ∼3 on the MRC dyspnoea grade).

**Table 1:**
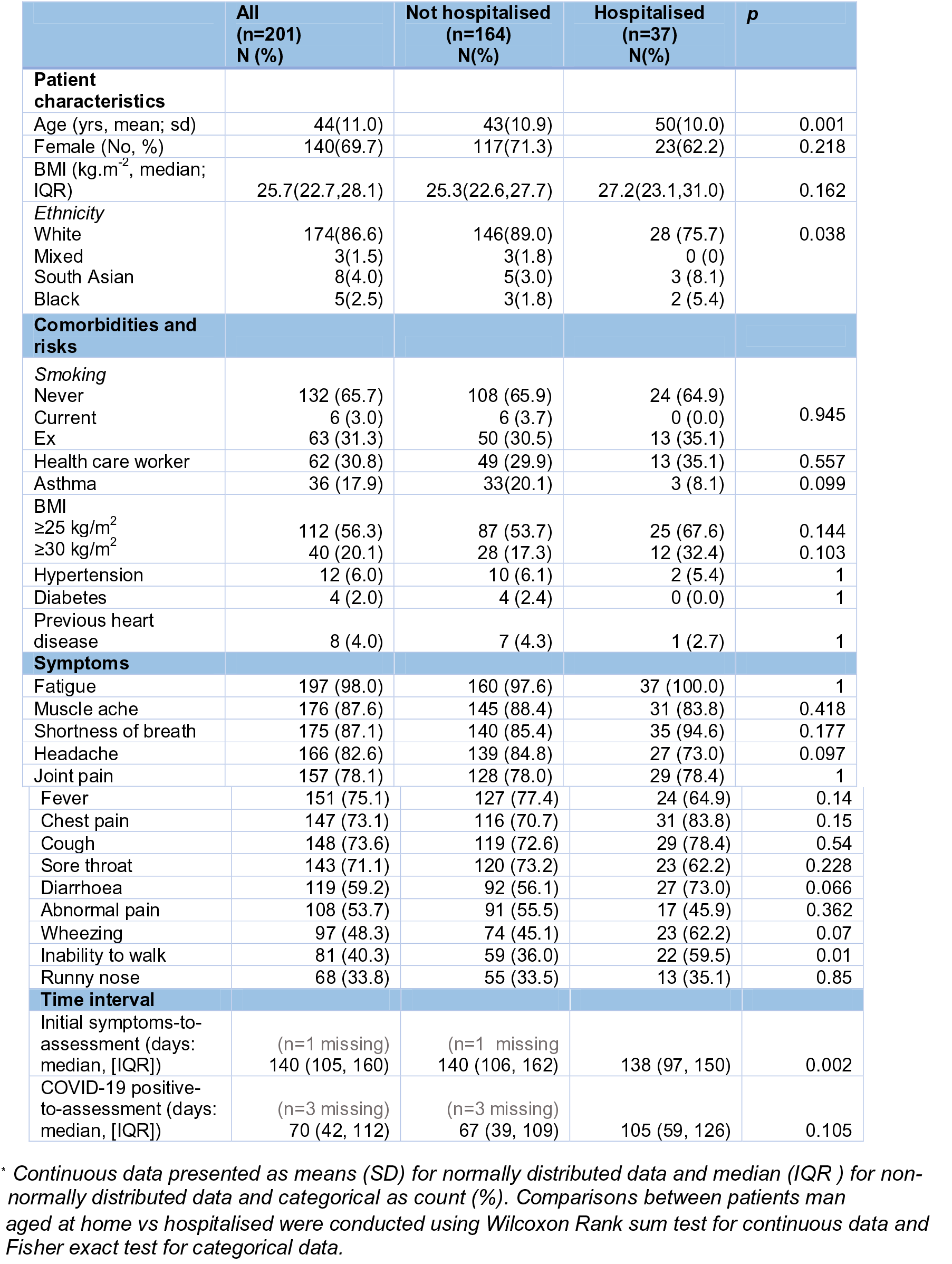
Baseline demographics and symptoms in 201 low-risk individuals with long-COVID.*

**Figure 2:**
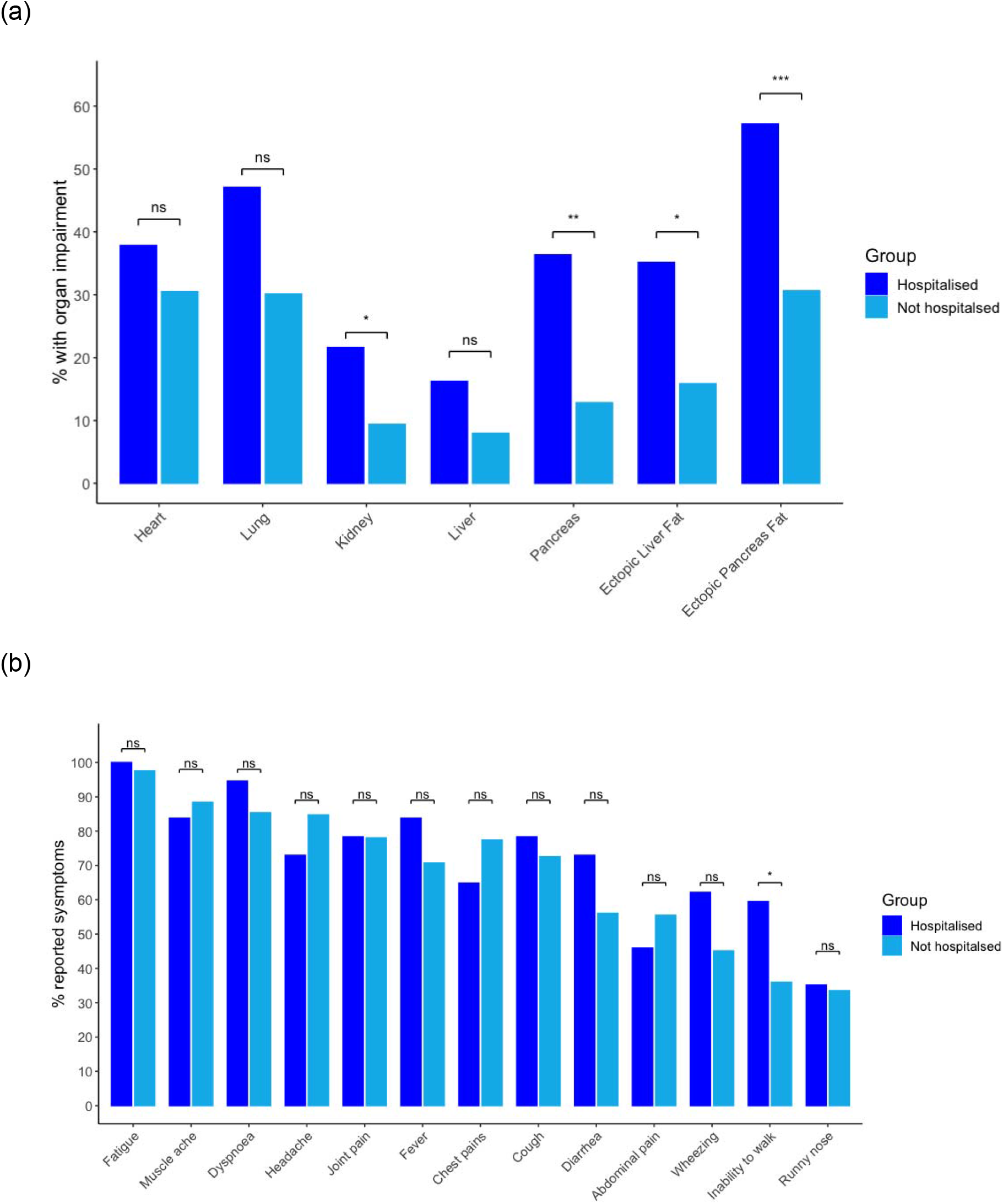
Proportion of low-risk individuals with long-COVID by hospitalisation (n=201) for (a) symptoms; and (b) evidence of organ impairment.

### Blood investigations

Triglycerides (p=0.002), cholesterol (p=0.021), LDL-cholesterol (p=0.005) and transferrin saturation (p=0.005) were more likely to be abnormal in hospitalised versus non-hospitalised individuals. Mean corpuscular haemoglobin concentration (26%), alanine transferase (14%), lactate dehydrogenase (16%), triglycerides (12%) and cholesterol (42%) were all abnormally high in ≥10% of all individuals (without separation by hospitalisation status). ESR (13%), bicarbonate (13%), uric acid (16%) and high-sensitivity CRP (13%) were abnormally high in in ≥10% of individuals in the hospitalisation group. Bicarbonate (10%), phosphate (13%), uric acid (11%), and transferrin saturation (19%) were abnormally low in ≥10% of individuals (without separation by hospitalisation status) (**Table S2**).

### Single and multi-organ impairment

Impairment was present in the heart in 32% (myocarditis in 11%; systolic dysfunction in 23%), lungs in 33%, kidneys in 12%, liver in 10%, pancreas in 17%, and 6% had evidence of splenomegaly (**Table 2, Figure 2(b)**). 66% of individuals had impairment in one or more organ systems. There was evidence of multi-organ impairment in 25% of individuals, with varying degrees of overlap across multiple organs (**Figure 1 and 3**). Organ impairment was more common in hospitalised versus non-hospitalised individuals. Measures of inflammation in the kidneys and pancreas, and ectopic fat in the pancreas and liver, were also higher in hospitalised individuals (all p <0.05)(**Figure 2(b)**).

**Table 2:** Evidence of organ impairment in 201 low-risk individuals with long-COVID.

| Measurement | All<br>(n=201)<br>N(%) | Not<br>hospitalised<br>(n=164)<br>N(%) | Hospitalised<br>(n=37)<br>N(%) | p |
| --- | --- | --- | --- | --- |
| HEART |  |  |  |  |
| Left ventricular ejection fraction (%) |  |  |  |  |
| • Normal (>55%) | 155 (77.1) | 129 (78.7) | 26 (70.3) | 0.079 |
| • Borderline impairment (50-55%) | 38 (18.9) | 31 (18.9) | 7 (18.9) |  |
| • Definite impairment (<50%) | 8 (4.0) | 4 (2.4) | 4 (10.8) |  |
| Left ventricular end diastolic volume (ml) |  |  |  |  |
| • >214ml in M; >178ml in W | 27 (13.4) | 18 (11.0) | 9 (24.3) | 0.057 |
| Evidence of myocarditis |  |  |  |  |
| • ≥ 3 segments with high T1 (≥1264ms at 3T; ≥1015ms at 1.5T) | 22 (10.9) | 18 (11.0) | 4 (10.8) | 1 |
| LUNGS |  |  |  |  |
| Deep Breathing Fractional area change | (n= 11 missing) | (n= 8 missing) | (n= 3 missing) |  |
| • < 39% | 63 (33.2) | 47 (30.1) | 16 (47.1) | 0.071 |
| KIDNEYS |  |  |  |  |
| Kidney cortex T1 | (n= 12 missing) | (n= 8 missing) | (n= 4 missing) |  |
| • Normal (<1610 ms at 3T; <1191ms at 1.5T) | 175 (88.4) | 146 (90.7) | 29 (78.4) | 0.046 |
| • Definite impairment (≥1610ms at 3T; ≥1191ms at 1.5T ) | 23 (11.6) | 15 (9.3) | 8 (21.6) |  |
| PANCREAS |  |  |  |  |
| Pancreatic inflammation (T1 in ms) |  |  |  |  |
| • Normal (<800ms) | 157 (83.1) | 136 (87.2) | 21 (63.6) | 0.003 |
| • Borderline (800-865ms) | 20 (10.6) | 11 (7.1) | 9 (27.3) |  |
| • Significant (>865ms) | 12 (6.3) | 9 (5.8) | 3 (9.1) |  |
| Pancreatic fat | (n= 6 missing) | (n= 4 missing) | (n= 2 missing) |  |
| • Normal (<5%) | 126 (64.6) | 111 (69.4) | 15 (42.9) | 0.005 |
| • Borderline (5-10%) | 44 (22.6) | 33 (20.6) | 11 (31.4) |  |
| • Significant(>10%) | 25 (12.8) | 16 (10.0) | 9 (25.7) |  |
| LIVER |  |  |  |  |
| Liver Inflammation (cT1 in ms) | (n= 1 missing) | (n= 1 missing) |  |  |
| • Normal (<800ms) | 181 (90.5) | 150 (92.0) | 31 (83.8) | 0.040 |
| • Borderline (800-825ms) | 5 (2.5) | 5 (3.1) | 0 (0.0) |  |
| • Significant (>825ms) | 14 (7.0) | 8 (4.9) | 6 (16.2) |  |
| Liver fat |  |  |  |  |
| • Normal (<5%) | 162 (80.6) | 138 (84.1) | 24 (64.9) | 0.025 |
| • Borderline (5-10%) | 18 (9.0) | 12 (7.3) | 6 (16.2) |  |
| • Definite (>10%) | 21 (10.4) | 14 (8.5) | 7 (18.9) |  |
| SPLEEN |  |  |  |  |
| Splenic length (mm) | (n= 10 missing) | (n= 10 missing) |  |  |
| • Normal (Table S1) | 179(9.4) | 144(9.5) | 35 (9.5) | 1 |
| • Borderline (Table S1) | 12 (6.3) | 10 (6.5) | 2 (5.4) |  |

\* Data are presented as count (%). Comparisons between patients managed at home vs hospitalised were conducted using Fisher exact test.

**Figure 3:**
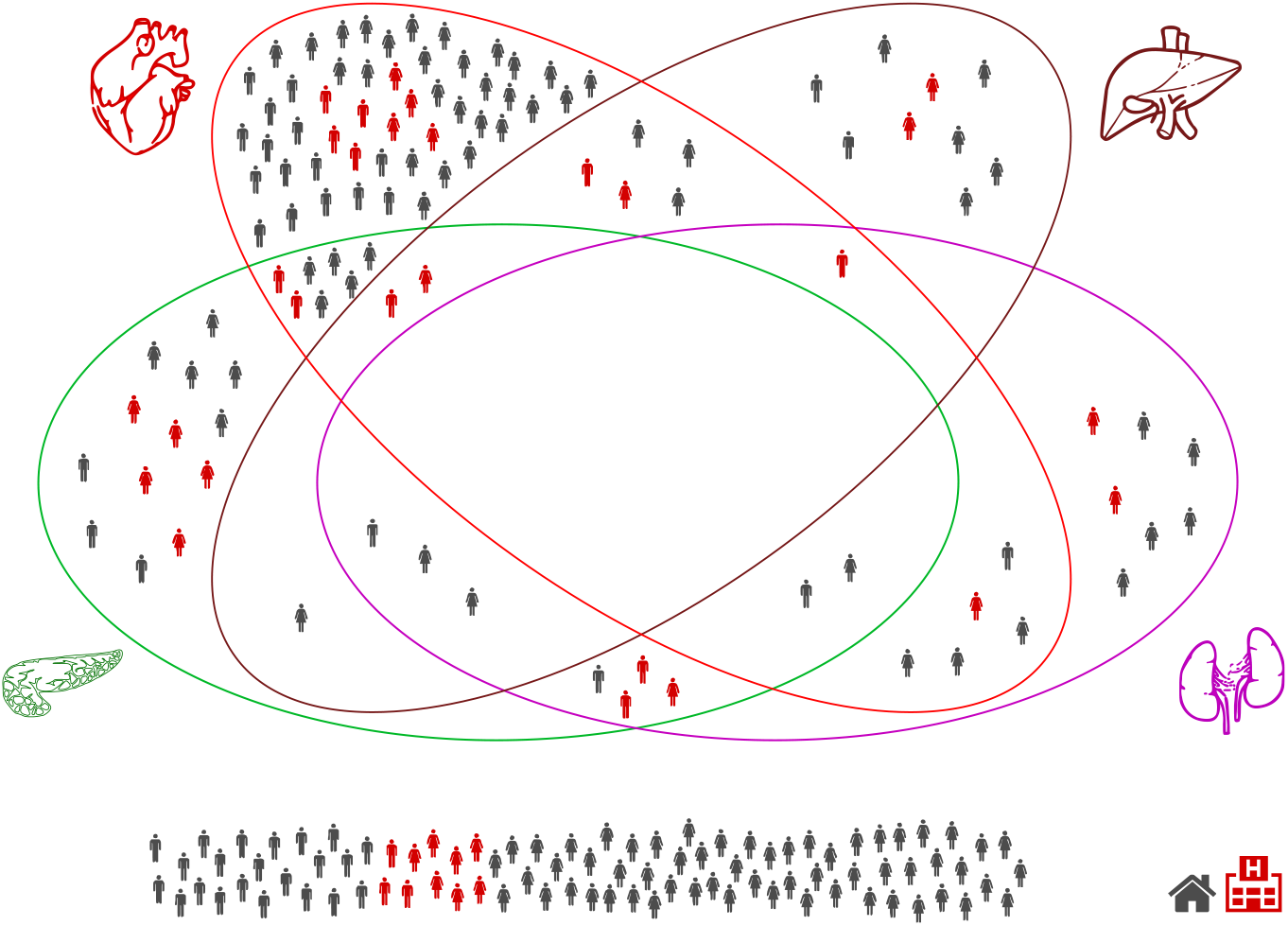
Multi-organ impairment in low-risk individuals with long COVID by gender and hospitalisation (n=201).

### Association between symptoms, blood investigations and organ impairment

**Figure 4** shows the percentage of reported symptoms in those with organ impairment (per organ). Multi-organ involvement was associated with more serious symptoms (fatigue, breathlessness etc), but no clear pattern was observed linking symptoms to organ impairment. Regression analysis did not show any association between specific organ impairment and specific symptoms or blood investigations. Increasing age (OR: 1.06 [CI: 1.02-1.10], p< 0.01), increased liver volume (OR: 1.18 [CI: 1.06-1.30], p<0.001) and having multi-organ impairment (OR: 2.75 [CI:1.22-6.22], p <0.05), all significantly increased the likelihood of being hospitalized, adjusting for gender and BMI.

**Figure 4:**
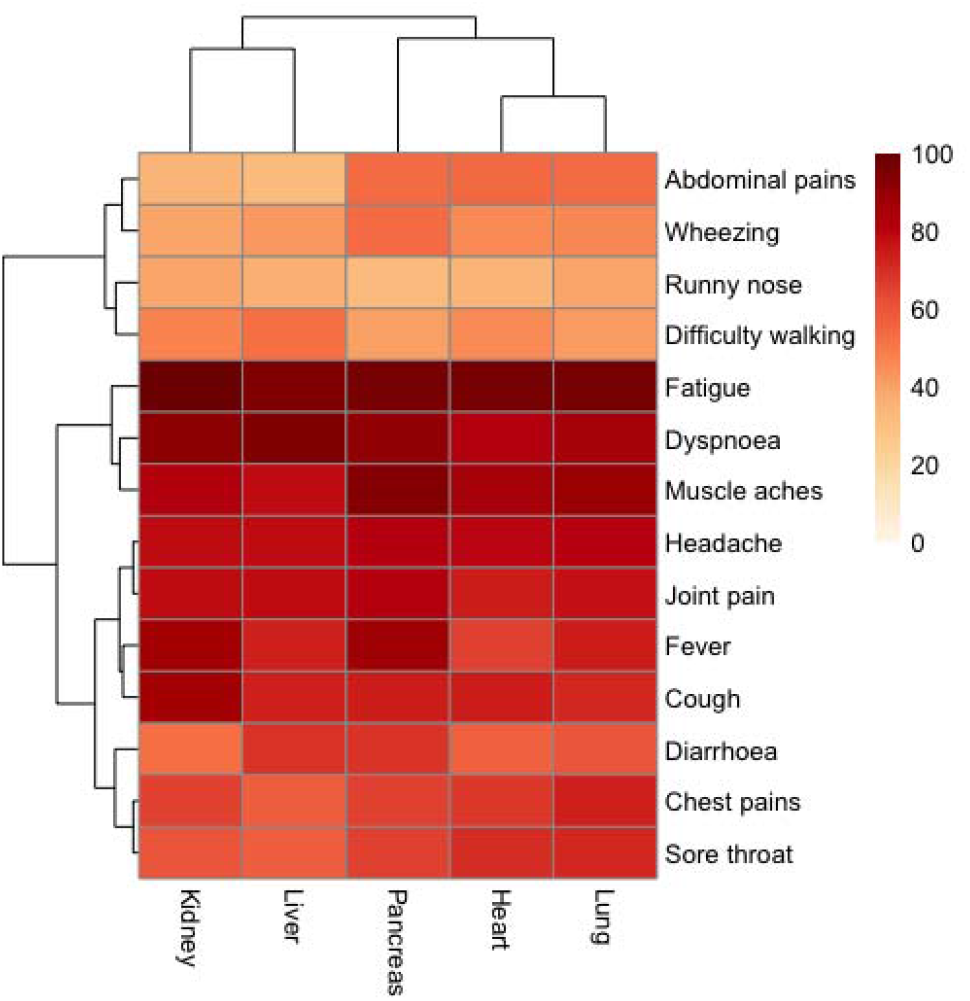
Clustering of reported symptoms and organ impairment for individuals with long-COVID (n=201).

## Discussion

In the first study to-date evaluating medium-term impairment across multiple organs following SARS-CoV2 infection, we had three major findings. First, in young individuals, largely without risk factors, pre-existing disease or hospitalisation, there was significant symptom burden and evidence of heart, lung, liver and pancreas impairment four months post-COVID-19. Second, symptoms and blood investigations predicted neither organ impairment nor hospitalisation. Third, cardiac (myocarditis and systolic dysfunction) and lung impairment have similar prevalence in low-risk individuals with long COVID.

The short-term symptoms likely to predict COVID-19(20) persist four months post-infection, particularly fatigue, shortness of breath, myalgia, headache and arthralgia. In this young cohort with low prevalence of comorbidities, the extent of symptom burden and organ impairment is concerning. Models of population COVID-19 impact have been based on age, underlying conditions and mortality, excluding morbidity or potential for multi-organ impairment and chronic diseases(21, 22). Moreover, studies highlighting extrapulmonary COVID-19 manifestations emphasised acute phase of illness(20). Although we describe mild rather than severe organ impairment, the pandemic’s scale and high infection rates in lower risk individuals (by age and underlying conditions), suggest a medium- and longer-term impact of SARS-CoV-2 infection which cannot be ignored in healthcare or policy spheres.

Although there may be an immunologic basis for variations in progression and severity of SARS-CoV-2 infection in different individuals(24), prediction models to-date have high rates of bias and poor performance(25). We found clustering of cardiorespiratory and gastrointestinal symptoms with evidence of impairment in heart, liver and pancreas respectively, but blood investigations were not associated with particular patterns of organ impairment as determined by COVERSCAN multi-organ assessment. Neither symptoms nor blood investigations were predictive of organ impairment. In acutely unwell patients, the focus has been on recognition of respiratory dysfunction and early provision of ventilatory support, but chronic multi-organ function has not been described systematically. Ongoing studies are considering chronic impact of COVID-19(26) but excluding non-hospitalised, low-risk individuals with and without organ impairment, which we will be investigating further in the longer term. As well as interest in specialist long COVID clinical services(27), there is a role for multi-organ assessment and ongoing evaluation, including low-risk, non-hospitalised individuals, perhaps even in the absence of symptoms.

Acute myocarditis and cardiogenic shock have been described(28), as well as high prevalence of myocarditis in hospitalised COVID-19 patients(29). In American athletes, although recent COVID-19 was associated with myocarditic changes, many non-infected patients also showed these changes(30). We now add that one third of low-risk individuals with long COVID syndrome have cardiac impairment in the form of mild systolic dysfunction or myocarditis three months following SARS-CoV-2 infection. Whilst causality cannot be attributed, cardiac function can be viewed as a risk factor for severe infection and an explanation of persistent symptoms in long COVID. As longitudinal data across organs become available, potential significance of our findings in the liver, kidney and pancreas needs to be explored.

### Implications for research

Our findings at four months post-infection and future findings have three research implications. First, as countries face second pandemic waves, models of the pandemic’s impact must include long COVID, whether quality of life, healthcare utilisation, productivity and economic effects. Second, there is urgent need for further multi-organ assessment, including blood and imaging analysis in the COVID-19 context, as well as linkage with primary and secondary care data, so that long COVID can be properly defined. Third, further longitudinal investigation of clustering of symptoms and organ impairment will inform health services research to plan multidisciplinary care pathways.

### Implications for clinical practice and public health

There are three implications for COVID-19 management. First, as well as highlighting the potential for MRI across organ systems following SARS-CoV-2 infection, our findings signal the need for monitoring and follow-up in at least the medium- and longer-term, especially for extrapulmonary sequelae. Second, as the search for effective COVID-19 vaccines and treatments continues, potential and real long-term multi-organ consequences of SARS-CoV-2 infection in low-risk individuals reinforce the central importance of minimising infection through social distancing, wearing of masks, physical isolation and other population-level measures. Third, both in terms of managing baseline risk, and monitoring and treating complications across organ systems, long COVID requires management across specialities (e.g. cardiology, gastroenterology) and disciplines (e.g. communicable and non-communicable diseases).

### Strengths and limitations

Our study is an ongoing, prospective, longitudinal cohort study with detailed blood and imaging characterisation of organ function, despite limited clinical examination with video consultations in the era of COVID-19. By recruiting ambulatory patients after infection with broad inclusion criteria (e.g. SARS-CoV-2 testing by virus RNA, antibody or antigen), we focus on individuals at lower risk of severity and mortality from acute SARS-CoV-2 infection. Our cardiac MRI protocol excluded gadolinium contrast as concerns regarding COVID-19-related renal complications remain. We relied on native T1 mapping to detect and characterise myocardial inflammation, allowing non-invasive tissue characterisation which was previously evaluated as superior to gadolinium MRI for acute myocarditis(31).

We report baseline findings following SARS-CoV-2 infection. In our pragmatic study design, the diagnosis of COVID-19 was by multiple methods, partly limited by access to laboratory testing during the pandemic. Causality of the relationship between organ impairment and infection cannot be deduced, but may be addressed by longitudinal follow-up of individuals with organ impairment. Our study population was limited by ethnicity despite disproportionate impact of COVID-19 in non-white individuals. Pulse oximetry and spirometry were added later to the protocol and follow up; they were not included from the outset to limit interaction and exposure between trial team and patients. We did not include healthy controls or MRI assessment of brain or muscle function.

## Conclusions

Long COVID has a physiological basis, with measurable patient-reported outcomes and organ impairment. Medium- and long-term evaluation and monitoring of multi-organ function beyond symptoms and blood investigations is likely to be required, even in lower risk individuals. Health system responses should emphasise suppression of population infection rates, as well as management of pre- and post-COVID-19 risk factors and chronic diseases.

## Data Availability

Data relating to this study is not available while the study is ongoing but will be made available by contacting the study team after recruitment and completion of follow-up.

## COVERSCAN study investigators

Perspectum: Mary Xu, Faezah Sanaei-Nezhad, Andrew Parks, Andrea Borghetto, Matthew D Robson, Petrus Jacobs, John Michael Brady, Carla Cascone, Soubera Rymell, Jacky Law, Virginia Woolgar, Velko Tonev, Claire Herlihy, Rob Suriano, Tom Waddell, Henrike Puchte, Alessandra Borlotti, Arun Jandor, Freddie Greatrex, Robin Jones, Georgina Pitts, Ashleigh West, Marion Maguire, Anu Chandra, Naomi Jayaratne, Dali Wu, Stella Kin, Mike Linsley, Valentina Carapella, Isobel Gordon, George Ralli, John McGonigle, Darryl McClymont, Boyan Ivanov, James Owler, Diogo Cunha, Tatiana Lim, Carlos Duncker, Madison Wagner, Marc Goldfinger, Adriana Roca, Charlotte Erpicum, Matthew David Kelly, Rexford D Newbould, Catherine J Kelly, Andrea Dennis, Sofia Mouchti, Arina Kazimianec, Helena Thomaides-Briers, Rajarshi Banerjee Mayo Clinic: Sandeep Kapur, Louise McLaughlin, Stacey A. Rizza University College London: Amitava Banerjee Great Western Hospitals NHS Foundation Trust: Malgorzata Wamil University of Oxford: Yi-Chun Wang, Tom Waddell

## Contributorship statement

Study design: AD, SK, RB, JA, SR

Patient recruitment: SK, RB, COVERSCAN team

Data collection: MW, LM, COVERSCAN team

Data analysis: AD, COVERSCAN team, AB

Data interpretation: AB, AD, MW, RB

Initial manuscript drafting: AB, AD, RB

Critical review of early and final versions of manuscript: all authors

Specialist input: cardiology (MW, AB); general medicine (RB, ADB, GAD); infectious disease (SAR, ADB); imaging (MR, RB); statistics (AD); epidemiology/public health (AB); primary care (SK); healthcare management (JA).

## Funding acknowledgements

AB is supported by research funding from NIHR, British Medical Association, Astra-Zeneca, UK Research and Innovation, and the Innovative Medicines Initiative-2 (BigData@Heart Consortium, under grant agreement No. 116074, supported by the European Union’s Horizon 2020 research and innovation programme and EFPIA; chaired by DE Grobbee and SD Anker, partnering with 20 academic and industry partners and ESC). This work was supported by the UK’s National Consortium of Intelligent Medical Imaging through the Industry Strategy Challenge Fund, Innovate UK Grant 104688, and also through the European Union’s Horizon 2020 research and innovation programme under grant agreement No 719445.

## Research in Context

### Evidence before this study

We searched PubMed, medRxiv, bioRxiv, arXiv, and Wellcome Open Research for peer-reviewed articles, preprints, and research reports on long COVID syndrome and medium- and long-term impact of coronavirus disease 2019 (COVID-19), using the search terms “coronavirus”, “COVID-19”, and similar terms, “organ impairment”, “organ function” and “morbidity”, up to September 30, 2020. We found no prior studies of medium- or long-term multi-organ impairment due to COVID-19. Prior studies have considered acute phase of illness and hospitalised patients, focusing on “high-risk” individuals based on age and underlying conditions. Without longer term data including lower risk individuals, full population impact of the pandemic cannot be assessed and health system responses cannot be planned.

### Added value of this study

In 201 individuals with low risk for COVID-19 severity and mortality (mean age 44 years, 20% obesity, 6% hypertension, 2% diabetes and 4% heart disease, 18% hospitalised), we assessed symptoms, blood investigations and multi-organ magnetic resonance imaging across organ systems, four months following SARS-CoV-2 infection. 99% and 42% had ≥4 and ≥10 symptoms respectively. Mild organ impairment was present in at least one organ in 66% and in 2 or more organs in 25% of individuals. Multi-organ impairment was associated with hospitalisation.

### Implications of all the available evidence

These analyses support strategies to suppress and minimise the infection rate in the population; medium- and long-term follow-up after SARS-CoV-2 infection with detailed evaluation across organ systems; and management of underlying conditions and risk factors before and after infection. For the first time, we provide multi-organ assessment in young, low-risk individuals with long COVID to inform healthcare and policy responses.

## Notes

### Competing Interest Statement

RB and AB are CEO and Head of Biomarker Science respectively at Perspectum. AB has received unrelated research grants from Astra Zeneca.

### Clinical Trial

NCT04369807

### Funding Statement

This work was supported by the UK National Consortium of Intelligent Medical Imaging through the Industry Strategy Challenge Fund, Innovate UK Grant 104688, and also through the European Union Horizon 2020 research and innovation programme under grant agreement No 719445.

### Author Declarations

Berkshire B ethics committee,UK (20/SC/0185)-ethical approval given.

