## Supplementary for "Multi-organ impairment in low-risk individuals with long COVID"

**Web Supplementary Materials**

**Supplementary methods	2**

**Supplementary references	4**

**Table S1: Reference ranges to define organ impairment	6**

**Table S2: Blood investigations in 201 low-risk individuals with long-COVID	8**

**Supplementary methods**

All the imaging methods can be deployed on standard clinical MRI scanners and are generally expedited approaches of methods previously demonstrated in the scientific literature that unless stated each utilise a short (<14seconds) breath-hold.

Cardiac imaging involved complete coverage of the heart with a short-axis stack (to the valve plane) of cine images acquired using cardiac gating, this acquisition mirrors that in UK Biobank and is a standardized approach(S1). Three short-axis cardiac T1 maps are acquired using the MOLLI-T1 approach at the basal, mid and apical levels of the left ventricle.

Liver and pancreas imaging used the LiverMultiScan acquisition protocol (Perspectum, Oxford, UK), which involves 3 single 2D axial slice breath-held acquisitions that separately are sensitive to the fat content (proton density fat fraction, or PDFF), to T2* (which is representative of liver iron content) and a MOLLI-T1 measurement (providing a measurement of tissue water), additionally a volumetric scan was used that covers the entire liver(S2).

Two dynamic cine MR acquisitions of the lung were acquired in the coronal plane with a 306.91 ms temporal resolution: one 40 s acquisition with the patient instructed to breathe normally and a second 30 s acquisition with the patient instructed to breathe deeply.

Kidney imaging used a single coronal view that was able to image both kidneys, imaging contrasts were MOLLI-T1, T2* (for blood oxygen level assessment), and diffusion imaging that was acquired during free-breathing in 2minutes.

**Image Analysis**

Cardiac MRI Analysis: Experienced cardiac MRI analysts used CVI42 (Cardiovascular Imaging Inc, Canada) to manually trace the end-diastolic and end-systolic phases in each of the short-axis views, following the standard UK BioBank evaluation approach as previously described(S3). This analysis yielded: For both the left and the right ventricle; End diastolic volume, End systolic volume, Stroke volume and Ejection Fraction. Additionally left ventricular muscle mass and wall thickness are determined from the function data. Cardiac T1 was determined for each of the 16 cardiac segments (of the AHA 17 segment model)(S4).

Liver Images were analysed by data analysts experienced at using the LiverMultiScan (Perspectum, Oxford, UK) software. This yielded global metrics in each liver of PDFF (proton density fat fraction), T2*, and cT1 (cT1 is a measurement of T1 that has been corrected for the confounding effects of iron and standardised to 3 Tesla; it is elevated with disease).

Pancreas images were analysed in a similar manner to the above except the software used was not FDA-cleared and iron correction was not performed. The output T1 was standardized to 3 Tesla.

Lung cine imaging allowed the measurement of the area of the left and right lungs through the breathing cycle in the coronal plane, which used automated methods that were reviewed by image analysts. The periodicity of the area fluctuations was used to determine the respiratory rate. All analysis was performed in-house using MATLAB based tools. The method was validated by measuring the correlation between the change in area and the forced vital capacity, the latter being measured using spirometry.

Kidney images were assessed using in-house tools to fit the parametric maps and allow trained analysts to make measurements. The T2* maps were analysed by the Twelve Layer Concentric Object (TLCO) approach that generates a gradient of relaxation values, in the other evaluations the cortex and medulla were manually segmented using the MOLLI-T1 map or the b=0 (in the case of diffusion) to guide the boundary.

In all cases the volumetric assessments utilised an initial in-house developed machine-learning driven segmentation, and then a manual step that may be used to fine tune boundaries. This approach was also used in the body composition analysis, which for reasons of speed was performed only in a single slice (an axial view that passes through L3 of the spine) in this work.

**Table S1: Reference ranges to define organ impairment**

|  | **Reference range** | **UK BioBank data** | **Rationale and evidence** | **On site healthy controls** |
| --- | --- | --- | --- | --- |
| **HEART** | | | | |
| Left ventricular ejection fraction (LVEF) | - Normal (>55%) - Borderline impairment (50-55%) - Definite impairment (<50%) | - | Meta-analysis: LVEF <57 %* in men and women aged 20-80 years (S5)  Guidelines: <50% Definite impairment (S6)  Observational data: Association of LVEF 50-55% with double the risk of HF over 12 years (HR: 2.15; 95% CI: 1.41 to 3.28) (S7) | - |
| Increased end-diastolic volume | - >214ml in men  - >178ml in women | - | Meta-analysis: Defined as upper limit in EDV (S5) | - |
| Myocarditis | More than 3 segments with abnormal T1. Abnormal T1 defined as:  - 1.5 Tesla: ≥1015 ms  - 3 Tesla: ≥1264 ms | - | - | T1 is a field-strength specific parameter in line with study-specific. Thresholds based on healthy controls in the same setting n=5. |
| **LUNGS** | | | | |
| Deep breathing fractional area change | Abnormal values are  < 39% | - | - | Thresholds based on healthy controls in the same setting n=9. |
| **LIVER** | | | | |
| Liver volume | Abnormal values are ≤2.13L | N=19489. Reference range defined by mean +2SD. Application 9914 | - | - |
| Liver fat | - Normal (<5%)  - Borderline fatty liver (5-10%)  - Definite fatty liver (>10%) | Normal has been defined as per UKBB Application 9914. (S9) N=4949 | Steatosis as described in NAFLD guidelines (S8). | - |
| Liver inflammation | - Normal (<800ms)  - Borderline (800-825ms)  - Significant (>825ms) | For the definition of Normal range (<721.9**) we used the upper limit for the range 40-49 years old. N=2816 from application 9914. (S9) | > 825ms predict liver-related clinical events, suggestive of significant fibroinflammation (S10) | - |
| **PANCREAS** | | | | |
| Pancreatic fat | - Normal (<5 %)  - Borderline (5- 10%)  - Significant (>10%) | Taken that this ranges are expressed in percentages; this values have been defined as per liver fat values. | - | - |
| Pancreatic inflammation | - Normal (<800ms)  - Borderline (8000-865ms)  - Significant (>865ms) | N=97 From application 9914. With BMI <25 and HbA1c <48  Normal ranges defined by mean +2SD and Significant impairment defined as mean +3SD | - | - |
| **KIDNEYS** | | | | |
| Renal Cortical T1 | - 1.5 Tesla: <1191ms  - 3 Tesla: <1610ms | - | Reference values based on healthy population; field strength dependent (S11-S12) | - |
| **SPLEEN** | | | | |
| Spleen volume | Abnormal values are ≤0.34L | N=19489. Reference range defined by mean +2SD | - | - |
| Spleen length | Abnormal values are:   - women   - ≥120mm for height <170cm   - ≥130mm for height ≥170cm - Men   - ≥130mm for height <170cm   - ≥135mm for height 175-185cm   - ≥140mm for height >185cm |  | Reference ranges defined as upper limit by gender and height (S13) | - |

***** Following expert medical advice, we have decided to set up normal as 55%, following the median of the UKBB values for Ejection fraction as per doctor´s recommendations. ****** Value has been rounded up to 800 ms

**Table S2: Blood investigations in 201 low-risk individuals with long-COVID**

Normal ranges for all the metrics are inclusive of values indicated on the low normal and high normal thresholds for each metric.

| **Measurement** | **All** | **Managed at home** | **Hospitalised** | **p-value** |
| --- | --- | --- | --- | --- |
| **Haemoglobin** |  |  |  |  |
| - Normal ( 130 - 170 g/L in men; 115 - 155 g/L in women ) | 160 (95.2%) | 131 (95.6%) | 29 (93.5%) | 0.588 |
| - Abnormal low ( < 130 g/L in men; < 115 g/L in women ) | 5 (3%) | 4 (2.9%) | 1 (3.2%) |  |
| - Abnormal high ( > 170 g/L in men; > 155 g/L in women ) | 3 (1.8%) | 2 (1.5%) | 1 (3.2%) |  |
| **Haematocrit (HCT)** |  |  |  |  |
| - Normal ( 0.37 - 0.5 in men; 0.33 - 0.45 in women ) | 163 (97%) | 133 (97.1%) | 30 (96.8%) | 0.396 |
| - Abnormal low ( < 0.37 in men; < 0.33 in women ) | 2 (1.2%) | 1 (0.7%) | 1 (3.2%) |  |
| - Abnormal high ( > 0.5 in men; > 0.45 in women ) | 3 (1.8%) | 3 (2.2%) | 0 (0%) |  |
| **Red cell count** |  |  |  |  |
| - Normal ( 4.4 - 5.8 x10^12/L in men; 3.95 - 5.15 x10^12/L in women ) | 160 (95.2%) | 131 (95.6%) | 29 (93.5%) | 0.299 |
| - Abnormal low ( < 4.4 x10^12/L in men; < 3.95 x10^12/L in women ) | 5 (3%) | 3 (2.2%) | 2 (6.5%) |  |
| - Abnormal high ( > 5.8 x10^12/L in men; > 5.15 x10^12/L in women ) | 3 (1.8%) | 3 (2.2%) | 0 (0%) |  |
| **Mean cell volume (MCV)** |  |  |  |  |
| - Normal ( 80 - 99 fL ) | 164 (97.6%) | 133 (97.1%) | 31 (100%) | 1 |
| - Abnormal low ( < 80 fL ) | 4 (2.4%) | 4 (2.9%) | 0 (0%) |  |
| - Abnormal high ( > 99 fL ) | 0 (0%) | 0 (0%) | 0 (0%) |  |
| **Mean corpuscular haemoglobin (MCH)** |  |  |  |  |
| - Normal ( 26 - 33.5 pg ) | 164 (97.6%) | 134 (97.8%) | 30 (96.8%) | 0.257 |
| - Abnormal low ( < 26 pg ) | 3 (1.8%) | 3 (2.2%) | 0 (0%) |  |
| - Abnormal high ( > 33.5 pg ) | 1 (0.6%) | 0 (0%) | 1 (3.2%) |  |
| **Mean corpuscular haemoglobin concentration (MCHC)** |  |  |  |  |
| - Normal ( 300 - 350 g/L ) | 125 (74.4%) | 100 (73%) | 25 (80.6%) | 0.496 |
| - Abnormal low ( < 300 g/L ) | 0 (0%) | 0 (0%) | 0 (0%) |  |
| - Abnormal high ( > 350 g/L ) | 43 (25.6%) | 37 (27%) | 6 (19.4%) |  |
| **Red cell distribution width (RDW)** |  |  |  |  |
| - Normal ( 11.5 - 15 ) | 151 (90.4%) | 120 (88.2%) | 31 (100%) | 0.224 |
| - Abnormal low ( < 11.5 ) | 10 (6%) | 10 (7.4%) | 0 (0%) |  |
| - Abnormal high ( > 15 ) | 6 (3.6%) | 6 (4.4%) | 0 (0%) |  |
| **Platelet count** |  |  |  |  |
| - Normal ( 150 - 400 x10^9/L ) | 157 (93.5%) | 130 (94.9%) | 27 (87.1%) | 0.258 |
| - Abnormal low ( < 150 x10^9/L ) | 1 (0.6%) | 1 (0.7%) | 0 (0%) |  |
| - Abnormal high ( > 400 x10^9/L ) | 10 (6%) | 6 (4.4%) | 4 (12.9%) |  |
| **Mean platelet volume (MPV)** |  |  |  |  |
| - Normal ( 7 - 13 fL ) | 167 (99.4%) | 136 (99.3%) | 31 (100%) | 1 |
| - Abnormal low ( < 7 fL ) | 0 (0%) | 0 (0%) | 0 (0%) |  |
| - Abnormal high ( > 13 fL ) | 1 (0.6%) | 1 (0.7%) | 0 (0%) |  |
| **White cell count** |  |  |  |  |
| - Normal ( 3 - 10 x10^9/L ) | 162 (96.4%) | 131 (95.6%) | 31 (100%) | 0.594 |
| - Abnormal low ( < 3 x10^9/L ) | 0 (0%) | 0 (0%) | 0 (0%) |  |
| - Abnormal high ( > 10 x10^9/L ) | 6 (3.6%) | 6 (4.4%) | 0 (0%) |  |
| **Neutrophils** |  |  |  |  |
| - Normal ( 2 - 7.5 x10^9/L ) | 155 (92.3%) | 125 (91.2%) | 30 (96.8%) | 0.832 |
| - Abnormal low ( < 2 x10^9/L ) | 10 (6%) | 9 (6.6%) | 1 (3.2%) |  |
| - Abnormal high ( > 7.5 x10^9/L ) | 3 (1.8%) | 3 (2.2%) | 0 (0%) |  |
| **Lymphocytes** |  |  |  |  |
| - Normal ( 1.2 - 3.65 x10^9/L ) | 153 (91.1%) | 123 (89.8%) | 30 (96.8%) | 0.309 |
| - Abnormal low ( < 1.2 x10^9/L ) | 15 (8.9%) | 14 (10.2%) | 1 (3.2%) |  |
| - Abnormal high ( > 3.65 x10^9/L ) | 0 (0%) | 0 (0%) | 0 (0%) |  |
| **Monocytes** |  |  |  |  |
| - Normal ( 0.2 - 1 x10^9/L ) | 167 (99.4%) | 136 (99.3%) | 31 (100%) | 1 |
| - Abnormal low ( < 0.2 x10^9/L ) | 0 (0%) | 0 (0%) | 0 (0%) |  |
| - Abnormal high ( > 1 x10^9/L ) | 1 (0.6%) | 1 (0.7%) | 0 (0%) |  |
| **Eosinophils** |  |  |  |  |
| - Normal ( 0 - 0.4 x10^9/L ) | 162 (96.4%) | 132 (96.4%) | 30 (96.8%) | 1 |
| - Abnormal high ( > 0.4 x10^9/L ) | 6 (3.6%) | 5 (3.6%) | 1 (3.2%) |  |
| **Basophils** |  |  |  |  |
| - Normal ( 0 - 0.1 x10^9/L ) | 168 (100%) | 137 (100%) | 31 (100%) | N/A |
| - Abnormal high ( > 0.1 x10^9/L ) | 0 (0%) | 0 (0%) | 0 (0%) |  |
| **Erythrocyte sedimentation rate (ESR)** |  |  |  |  |
| - Normal ( 1 - 20 mm/hr ) | 156 (91.8%) | 129 (92.8%) | 27 (87.1%) | 0.289 |
| - Abnormal low ( < 1 mm/hr ) | 0 (0%) | 0 (0%) | 0 (0%) |  |
| - Abnormal high ( > 20 mm/hr ) | 14 (8.2%) | 10 (7.2%) | 4 (12.9%) |  |
| **Sodium** |  |  |  |  |
| - Normal ( 135 - 145 mmol/L ) | 164 (97.6%) | 133 (97.1%) | 31 (100%) | 1 |
| - Abnormal low ( < 135 mmol/L ) | 3 (1.8%) | 3 (2.2%) | 0 (0%) |  |
| - Abnormal high ( > 145 mmol/L ) | 1 (0.6%) | 1 (0.7%) | 0 (0%) |  |
| **Chloride** |  |  |  |  |
| - Normal ( 98 - 107 mmol/L ) | 162 (96.4%) | 131 (95.6%) | 31 (100%) | 1 |
| - Abnormal low ( < 98 mmol/L ) | 4 (2.4%) | 4 (2.9%) | 0 (0%) |  |
| - Abnormal high ( > 107 mmol/L ) | 2 (1.2%) | 2 (1.5%) | 0 (0%) |  |
| **Bicarbonate** |  |  |  |  |
| - Normal ( 22 - 29 mmol/L ) | 141 (83.9%) | 117 (85.4%) | 24 (77.4%) | 0.201 |
| - Abnormal low ( < 22 mmol/L ) | 17 (10.1%) | 14 (10.2%) | 3 (9.7%) |  |
| - Abnormal high ( > 29 mmol/L ) | 10 (6%) | 6 (4.4%) | 4 (12.9%) |  |
| **Urea** |  |  |  |  |
| - Normal ( 1.7 - 8.3 mmol/L ) | 168 (100%) | 137 (100%) | 31 (100%) | N/A |
| - Abnormal low ( < 1.7 mmol/L ) | 0 (0%) | 0 (0%) | 0 (0%) |  |
| - Abnormal high ( > 8.3 mmol/L ) | 0 (0%) | 0 (0%) | 0 (0%) |  |
| **Creatinine** |  |  |  |  |
| - Normal ( 66 - 112 umol/L in men; 49 - 92 umol/L in women ) | 151 (89.9%) | 125 (91.2%) | 26 (83.9%) | 0.224 |
| - Abnormal low ( < 66 umol/L in men; < 49 umol/L in women ) | 12 (7.1%) | 9 (6.6%) | 3 (9.7%) |  |
| - Abnormal high ( > 112 umol/L in men; > 92 umol/L in women ) | 5 (3%) | 3 (2.2%) | 2 (6.5%) |  |
| **Bilirubin** |  |  |  |  |
| - Normal ( 0 - 20 umol/L ) | 166 (98.8%) | 136 (99.3%) | 30 (96.8%) | 0.336 |
| - Abnormal high ( > 20 umol/L ) | 2 (1.2%) | 1 (0.7%) | 1 (3.2%) |  |
| **Alkaline phosphatase** |  |  |  |  |
| - Normal ( 40 - 129 IU/L in men; 35 - 104 IU/L in women ) | 158 (94%) | 128 (93.4%) | 30 (96.8%) | 0.167 |
| - Abnormal low ( < 40 IU/L in men; < 35 IU/L in women ) | 8 (4.8%) | 8 (5.8%) | 0 (0%) |  |
| - Abnormal high ( > 129 IU/L in men; > 104 IU/L in women ) | 2 (1.2%) | 1 (0.7%) | 1 (3.2%) |  |
| **Aspartate transferase** |  |  |  |  |
| - Normal ( 0 - 37 IU/L in men; 0 - 31 IU/L in women ) | 154 (93.3%) | 126 (94%) | 28 (90.3%) | 0.435 |
| - Abnormal high ( > 37 IU/L in men; > 31 IU/L in women ) | 11 (6.7%) | 8 (6%) | 3 (9.7%) |  |
| **Alanine transferase** |  |  |  |  |
| - Normal ( 10 - 50 IU/L in men; 10 - 35 IU/L in women ) | 142 (84.5%) | 117 (85.4%) | 25 (80.6%) | 0.602 |
| - Abnormal low ( < 10 IU/L in men; < 10 IU/L in women ) | 2 (1.2%) | 2 (1.5%) | 0 (0%) |  |
| - Abnormal high ( > 50 IU/L in men; > 35 IU/L in women ) | 24 (14.3%) | 18 (13.1%) | 6 (19.4%) |  |
| **Lactate dehydrogenase (LDH)** |  |  |  |  |
| - Normal ( 135 - 225 IU/L in men; 135 - 214 IU/L in women ) | 132 (79.5%) | 109 (80.7%) | 23 (74.2%) | 0.219 |
| - Abnormal low ( < 135 IU/L in men; < 135 IU/L in women ) | 6 (3.6%) | 6 (4.4%) | 0 (0%) |  |
| - Abnormal high ( > 225 IU/L in men; > 214 IU/L in women ) | 28 (16.9%) | 20 (14.8%) | 8 (25.8%) |  |
| **Creatinine kinase (CK)** |  |  |  |  |
| - Normal ( 38 - 204 IU/L in men; 26 - 140 IU/L in women ) | 153 (91.1%) | 123 (89.8%) | 30 (96.8%) | 0.645 |
| - Abnormal low ( < 38 IU/L in men; < 26 IU/L in women ) | 2 (1.2%) | 2 (1.5%) | 0 (0%) |  |
| - Abnormal high ( > 204 IU/L in men; > 140 IU/L in women ) | 13 (7.7%) | 12 (8.8%) | 1 (3.2%) |  |
| **Gamma glutamyl transferase** |  |  |  |  |
| - Normal ( 10 - 71 IU/L in men; 6 - 42 IU/L in women ) | 155 (92.3%) | 127 (92.7%) | 28 (90.3%) | 0.468 |
| - Abnormal low ( < 10 IU/L in men; < 6 IU/L in women ) | 4 (2.4%) | 4 (2.9%) | 0 (0%) |  |
| - Abnormal high ( > 71 IU/L in men; > 42 IU/L in women ) | 9 (5.4%) | 6 (4.4%) | 3 (9.7%) |  |
| **Total protein** |  |  |  |  |
| - Normal ( 63 - 83 g/L ) | 163 (97%) | 134 (97.8%) | 29 (93.5%) | 0.23 |
| - Abnormal low ( < 63 g/L ) | 3 (1.8%) | 2 (1.5%) | 1 (3.2%) |  |
| - Abnormal high ( > 83 g/L ) | 2 (1.2%) | 1 (0.7%) | 1 (3.2%) |  |
| **Albumin** |  |  |  |  |
| - Normal ( 34 - 50 g/L ) | 156 (92.9%) | 126 (92%) | 30 (96.8%) | 0.698 |
| - Abnormal low ( < 34 g/L ) | 0 (0%) | 0 (0%) | 0 (0%) |  |
| - Abnormal high ( > 50 g/L ) | 12 (7.1%) | 11 (8%) | 1 (3.2%) |  |
| **Globulin** |  |  |  |  |
| - Normal ( 19 - 35 g/L ) | 163 (97%) | 133 (97.1%) | 30 (96.8%) | 0.396 |
| - Abnormal low ( < 19 g/L ) | 3 (1.8%) | 3 (2.2%) | 0 (0%) |  |
| - Abnormal high ( > 35 g/L ) | 2 (1.2%) | 1 (0.7%) | 1 (3.2%) |  |
| **Calcium** |  |  |  |  |
| - Normal ( 2.2 - 2.6 mmol/L ) | 162 (96.4%) | 132 (96.4%) | 30 (96.8%) | 0.441 |
| - Abnormal low ( < 2.2 mmol/L ) | 2 (1.2%) | 1 (0.7%) | 1 (3.2%) |  |
| - Abnormal high ( > 2.6 mmol/L ) | 4 (2.4%) | 4 (2.9%) | 0 (0%) |  |
| **Magnesium** |  |  |  |  |
| - Normal ( 0.6 - 1 mmol/L ) | 166 (98.8%) | 135 (98.5%) | 31 (100%) | 1 |
| - Abnormal low ( < 0.6 mmol/L ) | 1 (0.6%) | 1 (0.7%) | 0 (0%) |  |
| - Abnormal high ( > 1 mmol/L ) | 1 (0.6%) | 1 (0.7%) | 0 (0%) |  |
| **Phosphate** |  |  |  |  |
| - Normal ( 0.87 - 1.45 mmol/L ) | 142 (84.5%) | 114 (83.2%) | 28 (90.3%) | 0.516 |
| - Abnormal low ( < 0.87 mmol/L ) | 21 (12.5%) | 19 (13.9%) | 2 (6.5%) |  |
| - Abnormal high ( > 1.45 mmol/L ) | 5 (3%) | 4 (2.9%) | 1 (3.2%) |  |
| **Uric acid** |  |  |  |  |
| - Normal ( 266 - 474 umol/L in men; 175 - 363 umol/L in women ) | 137 (81.5%) | 114 (83.2%) | 23 (74.2%) | 0.116 |
| - Abnormal low ( < 266 umol/L in men; < 175 umol/L in women ) | 19 (11.3%) | 16 (11.7%) | 3 (9.7%) |  |
| - Abnormal high ( > 474 umol/L in men; > 363 umol/L in women ) | 12 (7.1%) | 7 (5.1%) | 5 (16.1%) |  |
| **Fasting triglycerides** |  |  |  |  |
| - Normal (< 2.3 mmol/L ) | 142 (88.2%) | 122 (92.4%) | 20 (69%) | 0.002 |
| - Abnormal high ( > 2.3 mmol/L ) | 19 (11.8%) | 10 (7.6%) | 9 (31%) |  |
| **Fasting cholesterol** |  |  |  |  |
| - Normal (< 5 mmol/L ) | 94 (58.4%) | 83 (62.9%) | 11 (37.9%) | 0.021 |
| - Abnormal high ( > 5 mmol/L ) | 67 (41.6%) | 49 (37.1%) | 18 (62.1%) |  |
| **HDL cholesterol** |  |  |  |  |
| - Normal ( 0.9 - 1.5 mmol/L in men; 1.2 - 1.7 mmol/L in women ) | 102 (60.7%) | 83 (60.6%) | 19 (61.3%) | 0.057 |
| - Abnormal low ( < 0.9 mmol/L in men; < 1.2 mmol/L in women ) | 15 (8.9%) | 9 (6.6%) | 6 (19.4%) |  |
| - Abnormal high ( > 1.5 mmol/L in men; > 1.7 mmol/L in women ) | 51 (30.4%) | 45 (32.8%) | 6 (19.4%) |  |
| **LDL cholesterol** |  |  |  |  |
| - Normal (< 3 mmol/L ) | 107 (65.2%) | 95 (70.4%) | 12 (41.4%) | 0.005 |
| - Abnormal high ( > 3 mmol/L ) | 57 (34.8%) | 40 (29.6%) | 17 (58.6%) |  |
| **Iron** |  |  |  |  |
| - Normal ( 10.6 - 28.3 umol/L in men; 6.6 - 26 umol/L in women ) | 154 (91.7%) | 126 (92%) | 28 (90.3%) | 0.232 |
| - Abnormal low ( < 10.6 umol/L in men; < 6.6 umol/L in women ) | 4 (2.4%) | 2 (1.5%) | 2 (6.5%) |  |
| - Abnormal high ( > 28.3 umol/L in men; > 26 umol/L in women ) | 10 (6%) | 9 (6.6%) | 1 (3.2%) |  |
| **Total iron binding capacity (TIBC)** |  |  |  |  |
| - Normal ( 41 - 77 umol/L ) | 162 (97%) | 132 (97.1%) | 30 (96.8%) | 1 |
| - Abnormal low ( < 41 umol/L ) | 0 (0%) | 0 (0%) | 0 (0%) |  |
| - Abnormal high ( > 77 umol/L ) | 5 (3%) | 4 (2.9%) | 1 (3.2%) |  |
| **Transferrin saturation** |  |  |  |  |
| - Normal ( 20 - 55 % ) | 132 (79%) | 114 (83.8%) | 18 (58.1%) | 0.005 |
| - Abnormal low ( < 20 % ) | 31 (18.6%) | 19 (14%) | 12 (38.7%) |  |
| - Abnormal high ( > 55 % ) | 4 (2.4%) | 3 (2.2%) | 1 (3.2%) |  |
| **High sensitivity CRP** |  |  |  |  |
| - Normal ( 0 - 5 mg/L ) | 135 (92.5%) | 114 (93.4%) | 21 (87.5%) | 0.39 |
| - Abnormal high ( > 5 mg/L ) | 11 (7.5%) | 8 (6.6%) | 3 (12.5%) |  |
